# Single-Nephron Dynamics Across Chronic Kidney Disease Stages in Overt Diabetic Nephropathy

**DOI:** 10.64898/2026.04.21.26351385

**Authors:** Akane Miura, Masahiro Okabe, Yusuke Okabayashi, Takaya Sasaki, Kotaro Haruhara, Nobuo Tsuboi, Takashi Yokoo

## Abstract

**Background:** Single-nephron glomerular filtration rate (GFR) represents a nephron-level functional index that may reveal key pathophysiological mechanisms driving progression in patients with diabetic nephropathy. However, its clinical relevance remains incompletely understood. This cross-sectional study assessed single-nephron estimated GFR (eGFR) across different chronic kidney disease (CKD) stages in patients with advanced diabetic nephropathy.

**Methods:** Nephron number was estimated as the number of nonglobally sclerotic glomeruli per kidney using computed tomography-derived cortical volume combined with biopsy stereology. Single-nephron eGFR was calculated by dividing eGFR by the nephron number of both kidneys. Patients were stratified according to CKD stage at kidney biopsy. Associations between CKD stages and single-nephron eGFR were evaluated using multivariable linear regression models adjusted for age, sex, urinary protein excretion, and eGFR.

**Results:** The study included 105 patients with biopsy-proven diabetic nephropathy and overt proteinuria (median age 59 years, 83% male, HbA1c 6.6%, 57% had nephrotic range proteinuria). The percentage of globally sclerotic glomeruli, mesangial expansion score, and prevalence of nodular lesions increased significantly with advancing CKD stage. Median nephron number declined from 529,178 to 224,458 per kidney, whereas glomerular volume remained constant. Single-nephron eGFR decreased markedly with CKD stage and remained significantly inversely associated with CKD stage after adjustment for clinicopathologic covariates (*P* for trend <0.001).

**Conclusion:** In overt diabetic nephropathy, single-nephron eGFR decreased with advancing CKD stage, despite relatively preserved glomerular volume. At this stage of disease, structural alterations specific to diabetic nephropathy may impair effective single-nephron filtration capacity.

## Introduction

Diabetic nephropathy is the leading cause of kidney failure worldwide^1^. The natural course of diabetic nephropathy typically unfolds over decades, with overt proteinuria often developing 10–15 years after the onset of diabetes and progressive kidney function decline occurring thereafter^2–4^. Dynamic changes in the whole-kidney glomerular filtration rate (GFR) are a hallmark of progressive diabetic nephropathy. The typical clinical course of progressive diabetic nephropathy is a transient increase in GFR in the early stage, followed by gradual progression from microalbuminuria to overt proteinuria^5^. Histopathologically, diabetic nephropathy is characterized by arteriolar hyalinosis and a spectrum of glomerular lesions, including diffuse mesangial expansion, exudative lesions, nodular sclerosis, and mesangiolysis/microaneurysm formation^6^.

In 1982, Brenner et al. proposed the hyperfiltration theory as a mechanism underlying the progression of chronic kidney disease (CKD), based on animal studies using micropuncture techniques^7^. According to this theory, a reduction in nephron number induces a compensatory increase in single-nephron GFR, leading to hyperperfusion injury and further nephron loss. Although this concept has shaped our understanding of CKD progression, including progression in diabetic nephropathy, the underlying mechanisms remain incompletely understood. In particular, the transition from compensatory single-nephron hyperfiltration to a progressive decline in whole-kidney function has not been fully elucidated.

Recently, Denic et al. established a method for estimating the total nephron number and single-nephron GFR in living humans using contrast-enhanced computed tomography (CT) images and kidney biopsies^8^. These methodological advances made it possible to calculate single-nephron GFR by dividing whole-kidney GFR by the estimated number of functioning nephrons. Our group further established a modified method for estimating kidney cortical volume using unenhanced CT images in living humans^9^. This method has enabled the estimation of total nephron number and single-nephron GFR in patients with kidney diseases, for whom contrast agents are often contraindicated.

Previous studies have reported associations between advancing CKD stage and histopathological findings in overt diabetic nephropathy^10–12^. However, how nephron-level parameters change across CKD stages in relation to the progression of these histopathological lesions remains unexplored. This gap is important, as nephron-level functional decline may represent a final common pathway linking structural injury to loss of whole-kidney GFR. The present cross-sectional study therefore aimed to estimate nephron-level parameters, including nephron number, glomerular volume, and single-nephron GFR, in patients with biopsy-proven diabetic nephropathy and to evaluate differences across CKD stages together with associated histopathological changes.

## Materials and Methods

### Patient selection

This study included adult Japanese patients with biopsy-proven diabetic nephropathy at The Jikei University Hospital (Tokyo), The Jikei University West Medical Center (Tokyo), and The Jikei University Kashiwa Hospital (Chiba, Japan) between 2007 and 2024. Indications for biopsy were diabetes with kidney dysfunction (eGFR <60 mL/min/1.73 m^2^) and/or persistent proteinuria of ≥0.5 g/day. Patients were retrospectively identified and excluded according to the following criteria: (i) CT images were not available within 1 year prior to kidney biopsy; (ii) kidney biopsy specimens contained fewer than 5 non-sclerotic glomeruli on light microscopy or a cortical area of less than 2 mm^2^; or (iii) there was histological evidence of glomerulopathy other than diabetic nephropathy. This study was approved by the Ethics Committee of the Jikei University School of Medicine for Biomedical Research (36-053 [12152]) and conducted according to the Declaration of Helsinki. In this retrospective cross-sectional study, participants were given the opportunity to opt out instead of providing informed consent.

### Definitions

Hypertension was defined as systolic blood pressure ≥140 mmHg, diastolic blood pressure ≥90mmHg, or the use of antihypertensive medications. Body surface area was determined by the following equation^13^: body surface area (m^2^) = Weight^0.425^ (kg) × Height^0.725^ (cm) × 71.84 × 10^−4^. The estimated GFR (eGFR) was calculated from serum creatinine using a modified equation for GFR based on Japanese individuals^14^: eGFR = 194 × age^−0.287^ × (serum creatinine level) ^−1.094^ (× 0.739 if female). CKD stages were defined based on eGFR and were classified into five categories: CKD G1–2, ≥60; G3a, 45 to <60; G3b, 30 to <45; G4, 15 to <30; and G5, <15 mL/min/1.73 m2. Urinary protein excretion (g/day) was measured using 24-hour urine collection. Nephrotic-range proteinuria was defined as urinary protein excretion of ≥3.5 g/day. Microhematuria was defined as ≥5 red blood cells per high-power field in urinary sediment.

### Histopathological analysis

All kidney tissue specimens were obtained by percutaneous biopsy using an 18-gauge needle. The tissues were embedded in paraffin, cut into 3 µm sections, and stained with hematoxylin-eosin, periodic acid-Schiff, Masson’s trichrome, and periodic acid silver-methenamine. All biopsy samples were stained by immunohistochemistry or immunofluorescence for IgG, IgA, IgM, C3, and C1q. A globally sclerotic glomerulus was defined as a glomerulus showing scarring lesions or hyaline deposition in more than 50% of its area. A nonglobally sclerotic glomerulus was defined as a glomerulus that was not globally sclerotic. Glomeruli containing segmental sclerosis were included among nonglobally sclerotic glomeruli. The areas of interstitial fibrosis/tubular atrophy and interstitial inflammation were observed at ×100 under an optical microscope, and the mean values were calculated from the scores assigned to each area in 5% increments. Arteriolar hyalinosis and arteriosclerotic lesions were graded as previously described^15^. Arteriolar hyalinosis grades: 0, no hyalinosis; 1, one or more areas of partial arteriolar hyalinosis; 2, hyalinosis involving ≤50% of the arteriolar wall; 3, hyalinosis involving >50% of the arteriolar wall or penetrating hyalinosis. Arteriosclerosis grades: 0, no intimal thickening; 1, intimal thickening/media thickness <1; 2, intimal thickening/media thickness ≥1. A similar grading approach was used to evaluate diabetes-specific glomerular lesions such as mesangial expansion, nodular lesions, exudative lesions, and mesangiolysis/microaneurysm formation. These findings were scored as follows. Mesangial expansion: 0, normal or mild change; 1, mesangial expansion < capillary lumen; 2, mesangial expansion = capillary lumen; 3, mesangial expansion > capillary lumen. Nodular lesions: 0, not detected; 1, one or more lesions detected in biopsy specimens. Exudative lesions: 0, not detected; 1, one or more lesions detected in biopsy specimens. Mesangiolysis/microaneurysm formation: 0, not detected; 1, one or more lesions detected in biopsy specimens. The proportions of these lesions in each specimen were also calculated quantitatively. Histopathological findings were also classified into class □ through □ based on the diabetic nephropathy classification of the Renal Pathology Society^6^. Class □: mild mesangial expansion (□ a) or severe mesangial expansion (□ b), but without nodular lesions or globally sclerotic glomeruli in more than 50% of glomeruli. Class □: one or more nodular lesions without class □ changes. Class □: more than 50% globally sclerotic glomeruli.

### Morphological measurements

The thickness of the obtained CT images was 5.0 mm. Kidney parenchymal volumes were measured as previously described using software (ITK-SNAP version 3.6; University of Pennsylvania, Philadelphia, PA, www.itksnap.org) to semi-automatically segment the parenchymal images obtained from unenhanced CT images of both kidneys^16^. Estimated kidney cortical volumes were calculated using the following equation: estimated kidney cortical volume (cm^3^) = –1.3 (intercept) + 0.71 × kidney parenchymal volume (cm^3^)^9^. The individual areas of all glomerular capillary tufts and the total area of the obtained kidney cortex were measured using image analysis software (NDP. View2; Hamamatsu Photonics, Shizuoka, Japan). Glomerular area was defined as the mean area enclosed by the outer capillary loops of the tuft. Mean glomerular volume was calculated from the measured glomerular area as follows: Mean glomerular volume 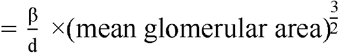, where β is a dimensionless shape coefficient (β = 1.382), and d is a size distribution coefficient (d = 1.01)^17^. The volumetric density of nonglobally sclerotic glomeruli was determined using the Weibel–Gomez stereological method as follows: nonglobally sclerotic glomerular density 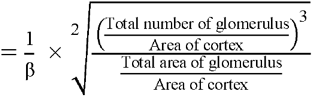, where β is a dimensionless shape coefficient (β = 1.38)17,18. Nephron number was defined as the number of nonglobally sclerotic glomeruli per kidney and was calculated as the product of the combined cortical volume of both kidneys and the volumetric density of nonglobally sclerotic glomeruli.^8^ The calculated value was then divided by 2 to obtain the value per kidney and multiplied by correction factors of 1.43 to account for tissue volume shrinkage due to paraffin embedding and 1.268 to account for volume shrinkage due to loss of tissue perfusion pressure. Single-nephron eGFR was calculated by dividing eGFR by the number of nonglobally sclerotic glomeruli in both kidneys.

### Statistical analysis

Patient characteristics at the time of kidney biopsy are presented as median (interquartile range [IQR]) for continuous variables, and as frequencies and proportions for categorical variables. The Mann–Whitney U test was used to compare continuous variables between two groups. The Jonckheere–Terpstra trend test was used to assess trends across ordered groups. For categorical variables, the Cochran–Armitage test was applied. To examine the independent association between CKD stage and nephron-level parameters, multivariable linear regression analyses were performed. Single-nephron eGFR was treated as the dependent variable, and CKD stage was entered as the independent variable. Models were adjusted for clinically relevant covariates, including age, sex, body surface area, urinary protein excretion, and use of renin–angiotensin–aldosterone system (RAAS) inhibitors. Given the right-skewed distribution of single-nephron eGFR and urinary protein excretion, natural logarithmic transformation was applied prior to regression modeling to approximate normality and stabilize variance. To assess the robustness of the findings, sensitivity analyses were performed in participant subgroups. Comparisons of nephron-level parameters among CKD stages were performed in patients whose biopsy specimens contained ≥10 glomeruli and a cortical area of ≥4 mm^2^, indicating adequate sampling, and in patients receiving renin-angiotensin-aldosterone system inhibitors at the time of kidney biopsy. All reported *P* values were two-sided, and *P* <0.05 was considered statistically significant. All statistical analyses were performed using R (version 4.2.1; The R Foundation for Statistical Computing, Vienna, Austria) with the R Commander interface, including the EZR extension (version 1.68; Saitama Medical Center, Jichi Medical University)^19^.

## Results

### Clinicopathological characteristics at the time of kidney biopsy

During the study period, 199 patients with biopsy-proven diabetic nephropathy were identified.

After excluding patients who met the exclusion criteria, 105 patients with diabetic nephropathy were included (**Figure 1**). A comparison of the clinical findings of patients included and excluded from the study is shown in **Supplementary Table S1**. Compared with excluded patients, included patients had a higher proportion of males, lower systolic blood pressure, higher HbA1c levels, and higher eGFR. No other significant differences were observed. The clinicopathological characteristics of the patients at the time of biopsy are shown in **Tables 1 and 2**. All patients included in the present study were Japanese, with a median age of 59 years, and 87 patients (83%) were male. All patients had already developed overt nephropathy both clinically and histopathologically, and 56 patients (57%) exhibited nephrotic-range proteinuria.

**Table 1.** Clinical characteristics and laboratory findings across CKD stages.

| Variables | All<br>(n = 105) | CKD stage |  |  |  |  | P value<br>for<br>trend |
| --- | --- | --- | --- | --- | --- | --- | --- |
|  |  | G1–2<br>(n = 15) | G3a<br>(n = 28) | G3b<br>(n = 30) | G4<br>(n = 19) | G5<br>(n = 13) |  |
| *Clinical findings* |  |  |  |  |  |  |  |
| Age (years) | 59 [46–69] | 50 [42–62] | 56 [48–70] | 60 [46–64] | 60 [51–70] | 67 [47–75] | 0.055 |
| Male; n (%) | 87 (83) | 11 (73) | 22 (79) | 25 (83) | 16 (84) | 13 (100) | 0.063 |
| BMI (kg/m<sup>2</sup>) | 24<br>[21–28] | 25<br>[21–30] | 25<br>[22–8] | 25<br>[22–29] | 24<br>[22–26] | 21<br>[21–26] | 0.13 |
| BSA (m2) | 1.7<br>[1.6–1.9] | 1.7<br>[1.5–1.9] | 1.8<br>[1.6–1.8] | 1.8<br>[1.7–1.9] | 1.7<br>[1.6–1.7] | 1.7<br>[1.5–1.7] | 0.26 |
| Patients with hypertension; n (%) | 97 (92) | 14 (93) | 25 (89) | 29 (97) | 17 (89) | 12 (92) | 0.99 |
| Systolic blood pressure (mmHg) | 137<br>[126–152] | 137<br>[130–143] | 133<br>[119–143] | 137<br>[130–157] | 135<br>[125–146] | 142<br>[138–161] | 0.10 |
| Diastolic blood pressure (mmHg) | 78 [70–87] | 80 [77–84] | 76 [66–85] | 75 [70–87] | 82 [70–85] | 82 [60–91] | 0.67 |
| RAAS inhibitor use; n (%) | 70 (67) | 10 (66) | 17 (61) | 22 (73) | 14 (73) | 7 (53) | 0.95 |
| *Treatment of diabetes* |  |  |  |  |  |  |  |
| Oral antidiabetic drug; n (%) | 57 (54) | 10 (66) | 19 (68) | 17 (56) | 15 (79) | 5 (38) | 0.33 |
| SGLT2 inhibitor use; n (%) | 14 (13) | 5 (33) | 3 (11) | 5 (17) | 1 (5) | 0 (0) | 0.069 |
| Insulin; n (%) | 18 (17) | 5 (33) | 4 (14) | 7 (23) | 6 (31) | 5 (38) | 0.33 |
| Oral antidiabetic drug + insulin; n (%) | 9 (9) | 3 (20) | 2 (7) | 1 (3) | 3 (16) | 0 (0) | 0.26 |
| *Laboratory findings* |  |  |  |  |  |  |  |
| Serum albumin (g/dL) | 2.9<br>[2.4–3.8] | 2.8<br>[2.4–4.0] | 3.2<br>[2.0–4.0] | 3.0<br>[2.2–3.3] | 3.3<br>[2.8–3.8] | 2.7<br>[2.5–2.9] | 0.55 |
| Serum creatinine (mg/dL) | 1.5<br>[1.0–2.1] | 0.8<br>[0.7–0.9] | 1.1<br>[1.0–1.2] | 1.6<br>[1.4–1.7] | 2.3<br>[2.0–3.0] | 5.6<br>[4.1–6.2] | < 0.001 |
| eGFR (ml/min/1.73m<sup>2</sup>) | 39<br>[26–53] | 72<br>[66–87] | 52<br>[48–54] | 36<br>[32–39] | 22<br>[18–26] | 8<br>[7–12] | < 0.001 |
| Serum uric acid (mg/dL) | 6.9<br>[5.9–7.9] | 7.0<br>[5.9–8.1] | 6.5<br>[5.8–7.0] | 6.4<br>[5.5–7.2] | 7.7<br>[7.0–8.3] | 8.5<br>[7.9–9.0] | < 0.001 |
| Triglyceride (mg/dL) | 165 [101– | 198 [116– | 150 [107– | 139 [106– | 165 [101– | 118 [96– | 0.24 |

|  | 252] | 300] | 239] | 294] | 225] | 247] |  |
| --- | --- | --- | --- | --- | --- | --- | --- |
| HDL cholesterol (mg/dL) | 51 [43–69] | 48 [44–61] | 56 [48–76] | 50 [40–64] | 48 [42–70] | 50 [41–67] | 0.49 |
| LDL cholesterol (mg/dL) | 113<br>[80–143] | 121<br>[79–145] | 121<br>[91–167] | 113<br>[81–133] | 103<br>[81–129] | 89<br>[64–139] | 0.19 |
| HbA1c (%) | 6.6<br>[5.9–7.3] | 7.1<br>[6.6–7.3] | 6.6<br>[6.1–7.1] | 6.3<br>[5.7–6.7] | 6.6<br>[5.9–7.5] | 6.8<br>[6.0–7.6] | 0.40 |
| Urinary protein excretion<br>(g/day) | 4.1<br>[1.6–6.3] | 4.3<br>[1.1–6.2] | 3.4<br>[1.2–6.2] | 4.3<br>[2.9–5.8] | 4.9<br>[2.7–9.4] | 3.0<br>[0.8–5.2] | 0.61 |
| Nephrotic range proteinuria; n<br>(%) | 60 (57) | 8 (53) | 14 (50) | 19 (63) | 13 (68) | 6 (46) | 0.69 |
| Microhematuria; n (%) | 40 (38) | 3 (20) | 10 (36) | 14 (47) | 9 (47) | 4 (30) | 0.32 |

Values are presented as the median [IQR1–3].
Abbreviations: BMI, body mass index; BSA, body surface area; CKD, chronic kidney disease; RAAS, renin-angiotensin aldosterone system; SGLT2, sodium-glucose cotransporter 2; eGFR, estimated glomerular filtration rate; HDL, high density lipoprotein; LDL, low density lipoprotein.

**Table 2.** Histopathological and morphometric findings across CKD stages.

| Variables | All<br>(n = 105) | CKD stage |  |  |  |  | P value for trend |
| --- | --- | --- | --- | --- | --- | --- | --- |
|  |  | G1–2<br>(n = 15) | G3a<br>(n = 28) | G3b<br>(n = 30) | G4<br>(n = 19) | G5<br>(n = 13) |  |
| *Histopathological findings* |  |  |  |  |  |  |  |
| Total number of glomeruli per biopsy | 24<br>[19–32] | 18<br>[13–24] | 25<br>[20–32] | 26<br>[23–36] | 26<br>[22–31] | 20<br>[19–24] | 0.31 |
| Glomeruli with global glomerulosclerosis (%) | 38<br>[22–54] | 19<br>[8–26] | 30<br>[19–42] | 44<br>[29–52] | 48<br>[38–58] | 60<br>[43–68] | < 0.001 |
| Glomeruli with segmental glomerulosclerosis (%) | 0 [0–5] | 0 [0–4] | 3 [0–8] | 0 [0–4] | 0 [0–6] | 0 [0–5] | 0.55 |
| Mesangial expansion, score 0–3 | 2.3<br>[2.0–2.8] | 2.5<br>[1.7–2.6] | 2.1<br>[1.8–2.2] | 2.5<br>[2.1–2.8] | 2.6<br>[2.3–2.9] | 2.6<br>[2.0–2.8] | 0.0034 |
| Exudative lesion; n (%) | 62 (59) | 8 (53) | 19 (68) | 22 (73) | 8 (42) | 5 (38) | 0.13 |
| Exudative lesion (%) | 7 [0–14] | 4 [0–8] | 8 [0–14] | 8 [1–14] | 0 [0–11] | 0 [0–14] | 0.69 |
| Nodular lesion; n (%) | 56 (53) | 4 (26) | 11 (39) | 21 (70) | 12 (63) | 8 (61) | 0.010 |
| Nodular lesion (%) | 6 [0–28] | 0 [0–3] | 0 [0–9] | 15 [0–29] | 16 [0–37] | 16 [0–33] | 0.0048 |
| Mesangiolytic/Microaneurysm; n (%) | 58 (55) | 5 (33) | 15 (53) | 21 (70) | 9 (47) | 8 (61) | 0.25 |
| Mesangiolytic/Microaneurysm (%) | 4 [0–20] | 0 [0–4] | 3 [0–14] | 11 [0–29] | 0 [0–15] | 6 [0–25] | 0.13 |
| Diabetic nephropathy classification |  |  |  |  |  |  |  |
| IIA; n (%) | 25 (24) | 5 (33) | 12 (43) | 5 (17) | 1 (5) | 2 (15) | 0.0092 |
| IIB; n (%) | 13 (12) | 6 (40) | 3 (11) | 2 (6) | 2 (11) | 0 (0) | 0.0058 |
| III; n (%) | 39 (37) | 4 (27) | 11 (35) | 14 (47) | 8 (42) | 3 (23) | 0.89 |
| IV; n (%) | 28 (27) | 0 (0) | 3 (11) | 9 (30) | 8 (42) | 8 (62) | < 0.001 |
| Interstitial fibrosis/tubular atrophy (%) | 57<br>[44–69] | 37<br>[32–47] | 47<br>[37–54] | 58<br>[44–71] | 68<br>[64–70] | 71<br>[70–80] | < 0.001 |
| Interstitial inflammation (%) | 30<br>[20–38] | 23<br>[15–31] | 21<br>[13–26] | 30<br>[19–42] | 37<br>[31–42] | 34<br>[31–42] | < 0.001 |
| Arteriolar hyalinosis, score 0–3 | 3.0<br>[2.0–3.0] | 3.0<br>[3.0–3.0] | 3.0<br>[2.7–3.0] | 3.0<br>[2.0–3.0] | 3.0<br>[2.5–3.0] | 3.0<br>[2.0–3.0] | 0.61 |
| Arteriosclerosis, score 0–2 | 1.0<br>[1.0–2.0] | 1.0<br>[1.0–2.0] | 1.0<br>[1.0–2.0] | 1.0<br>[1.0–2.0] | 1.0<br>[1.0–2.0] | 2.0<br>[1.0–2.0] | 0.26 |
| *Morphometric findings* |  |  |  |  |  |  |  |
| Kidney parenchymal volume<br>(cm<sup>3</sup>/kidney) | 153.6<br>[120.5–<br>185.4] | 187.7<br>[159.5–<br>223.3] | 158.0<br>[127.9–<br>181.7] | 152.0<br>[125.1–<br>173.1] | 120.8<br>[105.6–<br>175.4] | 129.4<br>[116.1–<br>158.2] | < 0.001 |
| Kidney cortical volume (cm<sup>3</sup>/kidney) | 109.0<br>[85.5–<br>131.6] | 133.2<br>[113.2–<br>158.5] | 112.2<br>[90.8–<br>129.0] | 107.9<br>[88.8–<br>122.9] | 85.8<br>[74.9–<br>124.5] | 91.9<br>[79.2–<br>112.3] | < 0.001 |
| Nonglobally sclerotic glomerular<br>density (per mm<sup>3</sup>) | 6.2<br>[4.9–8.5] | 7.6<br>[6.1–<br>12.2] | 7.1<br>[6.0–9.1] | 5.7<br>[4.9–7.6] | 5.7<br>[5.0–6.5] | 3.6<br>[3.0–5.1] | < 0.001 |
| Total number of nonglobally sclerotic<br>glomeruli<br>(× 10<sup>3</sup> per kidney) | 376 [262–<br>548] | 529 [464–<br>862] | 455 [326–<br>579] | 339 [270–<br>526] | 306 [221–<br>392] | 224 [138–<br>315] | < 0.001 |

Values are presented as the median [IQR1–3].

**Figure 1.**
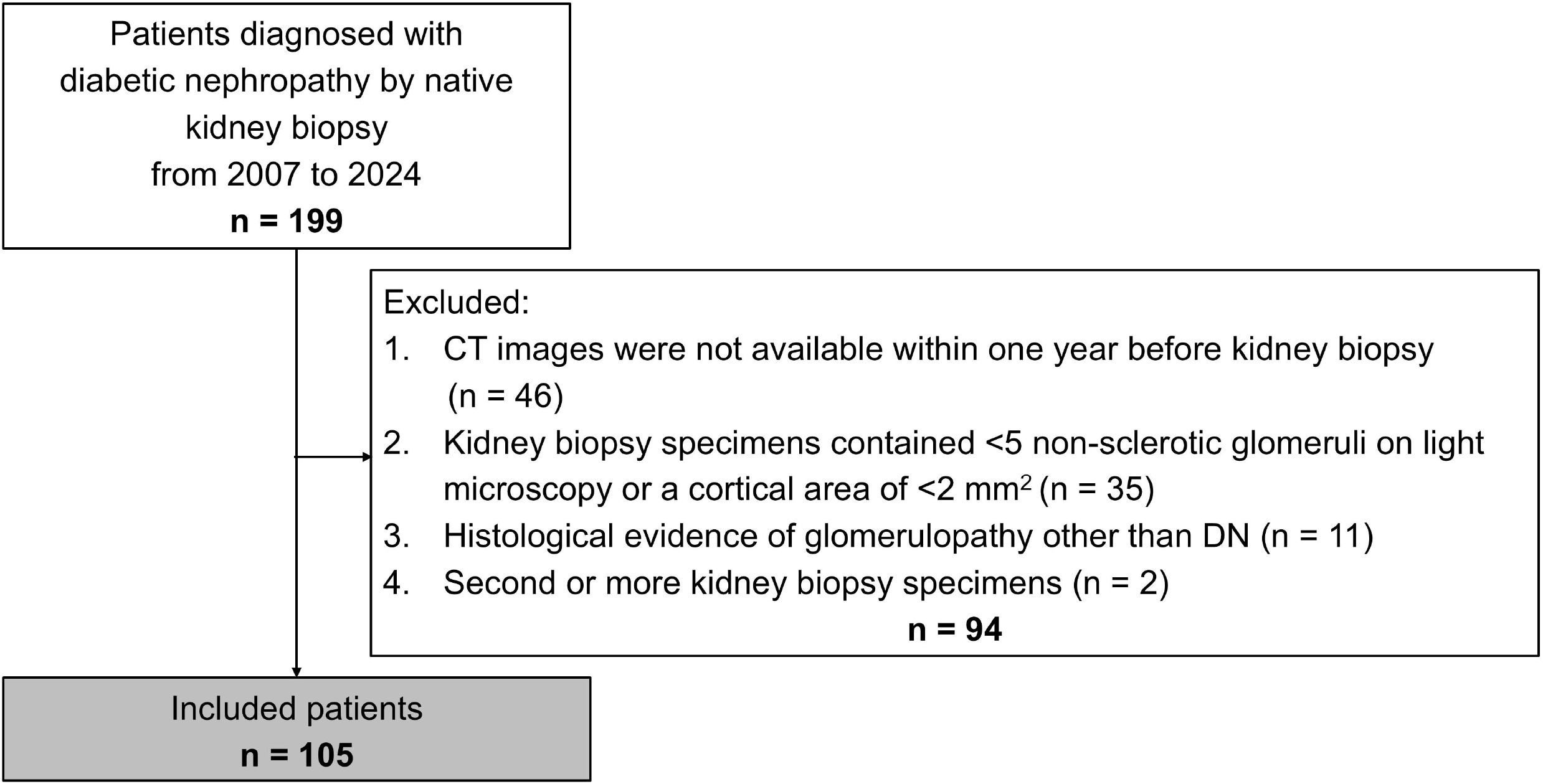
Patient selection. During the study period, 199 patients with biopsy-proven diabetic nephropathy were identified. Ninety-four patients were excluded from this study due to lack of CT data or insufficient glomerular number and/or cortical area in the biopsy specimens. CT, computed tomography.

### Comparison of clinicopathological findings across CKD stages

Comparison of clinicopathological findings across CKD stages are shown in **Tables 1** and **2**. Age increased with advancing CKD stage, but the trend was not statistically significant (*P* for trend = 0.055). BMI, systolic and diastolic blood pressure, HbA1c, and urinary protein excretion were not statistically different among CKD stages. As for the histopathological findings, the median percentage of globally sclerotic glomeruli, the mesangial expansion score, and the number of patients with nodular lesions were significantly higher with advancing CKD stages (*P* for trend = <0.001, 0.003, and 0.01, respectively). Other glomerular lesions specific to diabetic nephropathy were evident from early CKD stages, but there were no significant differences across CKD stages.

### Comparison of morphometric findings and single-nephron eGFR across CKD stages

Comparisons of morphometric findings and single-nephron eGFR between patients with different CKD stages are shown in **Table 2** and **Figure 2**. The measured parenchymal volume and the estimated cortical volume of the kidneys decreased with advancing CKD stages (*P* for trend < 0.001). As CKD stage advanced, nephron number, defined as the number of nonglobally sclerotic glomeruli, decreased significantly (*P* for trend < 0.001). While glomerular volume remained relatively constant across CKD stages, single-nephron eGFR significantly decreased with advancing CKD stages (*P* for trend < 0.001).

**Figure 2.**
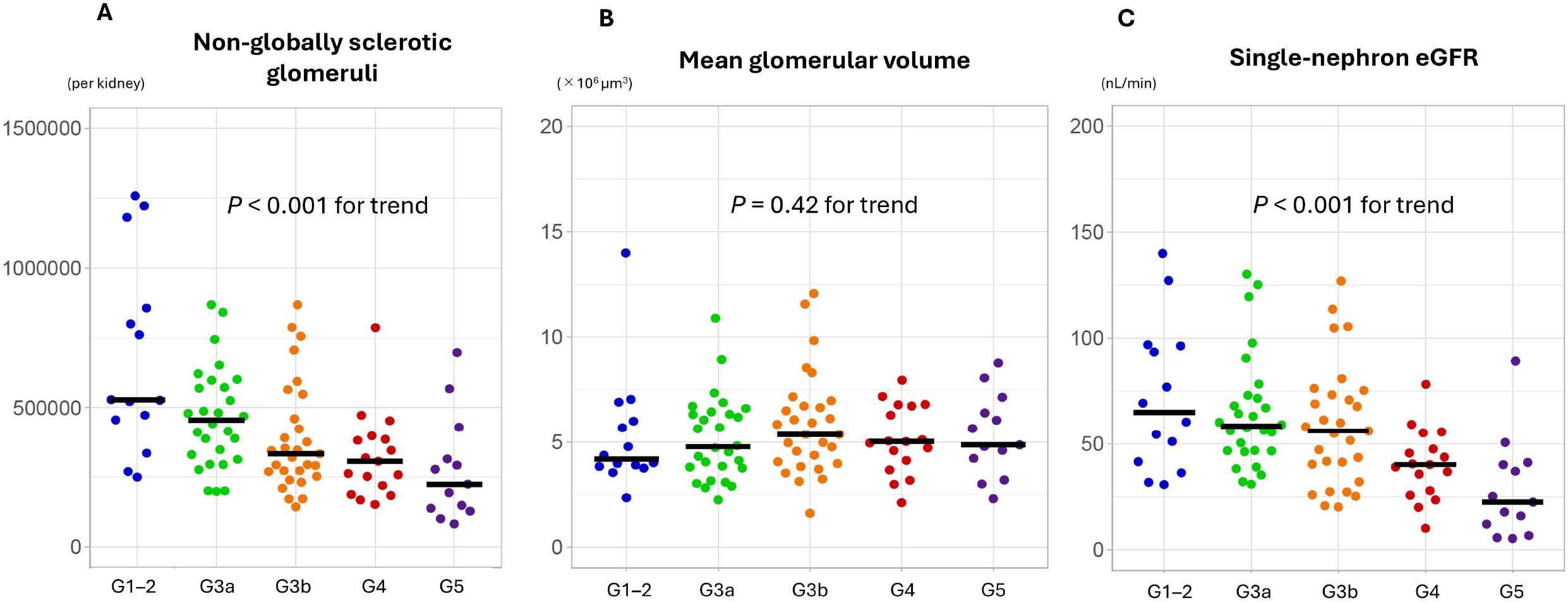
Comparison of nephron-level parameters across CKD stages. Nephron number per kidney (A), mean glomerular volume (B), and single-nephron eGFR (C) are shown at different CKD stages. As CKD stage advanced, nephron number per kidney decreased significantly. Although glomerular volume remained constant throughout CKD stages, single-nephron eGFR decreased significantly as CKD stage progressed. CKD, chronic kidney disease.

### Multivariable association between CKD stage and single-nephron eGFR

In multivariable linear regression analyses adjusted for age, sex, body surface area, urinary protein excretion, and use of RAAS inhibitors, single-nephron eGFR remained significantly associated with CKD stage (*P* for trend <0.001; **Table 3**). Compared with the combined CKD G1–2 group, adjusted single-nephron eGFR levels were significantly lower in patients with CKD stages G4 and G5.

**Table 3.** Multivariable adjusted single-nephron eGFR levels across CKD stages.

| CKD stage | Single-nephron eGFR (95% confidence interval), nL/min | *P* for trend |
| --- | --- | --- |
| G1–2 | 56.1 (42.8–73.5) | < 0.001 |
| G3a | 58.4 (48.2–70.8) |  |
| G3b | 52.3 (43.1–63.6) |  |
| G4 | 37.7 (29.7–48.0) * |  |
| G5 | 21.4 (15.8–28.8) * |  |

Models were adjusted for age, sex, body surface area, urinary protein excretion, and use of renin–angiotensin–aldosterone system inhibitors. Single-nephron eGFR and urinary protein excretion were logarithmically transformed.
\* *P* < 0.05 (vs. G1–2)
Abbreviations: CKD, chronic kidney disease; eGFR, estimated glomerular filtration rate.

### Sensitivity analyses

Sensitivity analyses were performed in patients whose biopsy specimens contained ≥10 glomeruli and a cortical area of ≥4 mm^2^, indicating adequate sampling (n = 78), and in patients treated with RAAS inhibitors, given their potential effects on single-nephron hemodynamics (n = 70). In each analysis, patients were categorized by CKD stage, and nephron-level parameters were compared. Nephron number, glomerular volume, and single-nephron eGFR showed trends consistent with the main findings of this study in both analyses (**Supplementary Figures S1** and **S2**).

## Discussion

In this cross-sectional study of patients with biopsy-proven diabetic nephropathy presenting with overt proteinuria, we evaluated differences in nephron-level parameters across CKD stages. As CKD stage advanced, nephron number declined, whereas glomerular volume remained unchanged. In contrast, single-nephron eGFR progressively decreased, indicating a dissociation between glomerular size and filtration capacity. This association between advancing CKD stage and lower single-nephron eGFR remained significant after adjustment for age, sex, body surface area, urinary protein excretion, and RAAS inhibitor use, suggesting that the decline in single-nephron eGFR is not solely attributable to systemic or hemodynamic factors. To our knowledge, this is the first study to systematically characterize nephron-level parameters in patients with diabetic nephropathy during the phase of progressive decline in kidney function.

Autopsy studies have shown that diabetic nephropathy-related structural lesions and glomerular enlargement are already present before the onset of overt albuminuria, particularly in individuals with both diabetes and hypertension^20–22^. In our cohort of patients with overt diabetic nephropathy, glomerular enlargement was already evident before an overt decline in kidney function, with mean glomerular volume in CKD stages G1–2 being nearly twofold greater than that observed in healthy kidney transplant donors^23^. This glomerular hypertrophy remained stable across CKD stages, whereas single-nephron eGFR declined progressively. These results suggest that glomerular hypertrophy had already reached a maximal plateau by the time diabetic nephropathy progressed to overt proteinuria. This may reflect a “ceiling effect” of structural remodeling, whereby the glomerulus remains enlarged while its internal filtration capacity progressively declines. This dissociation between sustained hypertrophy and declining single-nephron eGFR is consistent with a transition to a hypofiltration phase.

Brenner and colleagues described CKD progression using an animal model of progressive renal failure, in which experimental nephron reduction induces compensatory hypertrophy and increased single-nephron GFR, a response that ultimately becomes maladaptive and contributes to further nephron loss^24,25^. Consistent with this concept, nephron number in our cohort was already reduced even in CKD stages G1–2 (median 529,000 per kidney), markedly lower than that observed in healthy kidney transplant donors (median 626,000 per kidney)^23^. These findings indicate that a structural nephron deficit is already present at relatively early CKD stages in patients with overt diabetic nephropathy. This early structural deficit suggests that overt diabetic nephropathy may represent a clinical manifestation of the nephron reduction process described by Brenner and colleagues, in which nephron-level functional decline accompanies progressive kidney dysfunction.

Several reports have suggested that periglomerular arteriolar hyalinosis represents an earlylesion contributing to the development and progression of glomerular lesions^26,27^. However, in our cohort, nearly all patients (99%) exhibited periglomerular arteriolar hyalinosis, with no difference in its severity across CKD stages. These findings suggest that, in the overt phase of diabetic nephropathy, changes in single-nephron GFR are unlikely to be driven primarily by arteriolar hemodynamic factors. In line with this interpretation, previous biopsy studies in diabetic nephropathy patients proposed that a reduction in the effective glomerular capillary filtration surface area is a key mechanism of kidney function decline^28^. The discrepancy between glomerular size and filtration capacity observed in our study suggests that diabetic nephropathy-specific glomerular lesions such as mesangial expansion and nodular lesions act as “space-occupying” pathologies.

In multivariable analyses, the association between CKD stage and lower single-nephron eGFR remained significant after adjustment for age, sex, body surface area, urinary protein excretion, and use of RAAS inhibitors. Each of these factors may plausibly influence single-nephron hemodynamics or filtration capacity. Age may reflect cumulative structural injury^29^; body surface area relates to metabolic demand^30^; proteinuria may indicate ongoing glomerular stress^31^; and RAAS blockade can directly modify intraglomerular pressure^32^. Nevertheless, even after accounting for these potential influences, CKD stage remained independently associated with reduced single-nephron eGFR. This finding suggests that the decline in single-nephron eGFR is not merely a secondary consequence of systemic or treatment-related factors, but is intrinsically linked to the progression of structural injury that defines advancing CKD stage in overt diabetic nephropathy.

Some limitations merit consideration. First, the cross-sectional study design makes it difficult to infer causal relationships. Second, sampling bias due to needle biopsy cannot be ruled out. Although the results of the sensitivity analysis with only well-sampled patients were still robust, this bias should be considered when interpreting the results. Third, this study included only patients with advanced diabetic nephropathy who presented with overt proteinuria, not patients with early diabetic nephropathy. Therefore, this study cannot address changes in single-nephron eGFR in diabetic nephropathy in the early disease stages. Fourth, the number of patients treated with sodium-glucose cotransporter 2 inhibitors was very small, precluding analysis of their potential effects on single-nephron eGFR. Finally, all subjects in this study were Japanese, making it difficult to generalize the results to other races or geographic regions.

In conclusion, our study provides direct evidence that the progression of overt diabeticnephropathy is accompanied by a stage-dependent decline in single-nephron filtration capacity. Beyond the early hyperfiltration phase, advancing CKD stage is characterized by declining filtration function at the level of individual nephrons despite persistently enlarged glomeruli. These findings support the concept that the later course of diabetic nephropathy represents a hypofiltration phase emerging after years of sustained metabolic and hemodynamic stress. This highlights the central role of single-nephron dynamics in the progression of overt diabetic nephropathy.

## Acknowledgments

This study was approved by the Ethics Committee of the Jikei University School of Medicine for Biomedical Research [Approval No. 30-385 (9406)].

This work was supported by a Tokyo Diabetic Nephropathy Research Association Research Grant Fund to A.M. and a JSPS KAKENHI (Grant Number JP 25K11551) to N.T. The results of this study were presented at the American Society of Nephrology Kidney Week 2023.

## Data availability statement

All data produced in the present study are available upon reasonable request to the authors.

## Author contributions

N.T. conceptualized the study. A.M. and Y.O. performed measurements and data collection. A.M., M.O., T.S., and N.T. performed data analysis and drafted the manuscript. Y.O., K.H., and T.Y. provided critical intellectual input. All authors approved the final manuscript.

## Supplementary Data

**Supplementary Table S1**. Comparison of clinical characteristics between patients included in this study and those excluded from it.

**Supplementary Figure S1**. Comparison of nephron-level parameters across CKD stages in patients whose biopsy specimens contained ≥10 glomeruli and a cortical area of ≥4 mm^2^.

**Supplementary Figure S2**. Comparison of nephron-level parameters across CKD stages in patients treated with RAAS inhibitors.

