## Supplementary Data for "Single-Nephron Dynamics Across Chronic Kidney Disease Stages in Overt Diabetic Nephropathy"

**Supplementary Materials**

**Supplementary Table S1. Comparison of clinical characteristics between patients included in this study and those excluded from it.**

**Supplementary Figure S1. Comparison of nephron-level parameters across CKD stages in patients whose biopsy specimens contained ≥10 glomeruli and a cortical area of ≥4 mm^2^.**

**Supplementary Figure S2. Comparison of nephron-level parameters across CKD stages in patients treated with RAAS inhibitors.**

**Supplementary Table S1.** **Comparison of clinical characteristics between patients included in this study and those excluded from it.**

| Variables | All  (N = 105) | Excluded  (N = 94) | *P* value |
| --- | --- | --- | --- |
| *Clinical findings* |  |  |  |
| Age (years) | 59 [46–69] | 63 [51–70] | 0.18 |
| Male; n (%) | 87 (83) | 65 (69) | 0.030 |
| BMI (kg/m^2^) | 24.7 [21.8–28.3] | 24.2 [21.8–27.3] | 0.46 |
| Patients with hypertension; n (%) | 97 (92) | 79 (93) | 1 |
| Systolic blood pressure (mmHg) | 137 [126–152] | 144 [130–160] | 0.042 |
| Diastolic blood pressure (mmHg) | 78 [70–87] | 80 [70–88] | 0.44 |
| RAAS inhibitor use; n (%) | 70 (67) | 56 (66) | 1 |
| *Treatment of diabetes* |  |  |  |
| Oral antidiabetic drug; n (%) | 57 (54) | 44 (52) | 0.14 |
| SGLT2 inhibitor use; n (%) | 14 (13) | 5 (5) | 0.089 |
| Insulin; n (%) | 18 (17) | 28 (33) | 0.33 |
| Oral antidiabetic drug + insulin; n (%) | 9 (9) | 7 (8) | 1 |
| *Laboratory findings* |  |  |  |
| Serum albumin (g/dL) | 2.9 [2.4–3.8] | 2.9 [2.4–3.3] | 0.28 |
| Serum creatinine (mg/dL) | 1.5 [1.0–2.1] | 1.6 [1.2–2.9] | 0.072 |
| eGFR (ml/min/1.73m^2^) | 39 [26–53] | 29 [13–45] | 0.012 |
| Serum uric acid (mg/dL) | 6.9 [5.9–7.9] | 6.9 [5.9–8.2] | 0.96 |
| Triglyceride (mg/dL) | 165 [101–252] | 147 [109–184] | 0.25 |
| HDL cholesterol (mg/dL) | 51 [43–69] | 48 [43–69] | 0.71 |
| LDL cholesterol (mg/dL) | 113 [80–143] | 117 [89–142] | 0.73 |
| HbA1c (%) | 6.6 [5.9–7.3] | 6.1 [5.0–6.5] | <0.001 |
| Urinary protein excretion (g/day) | 4.1 [1.6–6.2] | 5.0 [1.9–7.5] | 0.26 |
| Nephrotic range proteinuria; n (%) | 60 (57) | 31 (38) | 0.55 |
| Microhematuria; n (%) * | 40 (38) | 28 (35) | 0.75 |

Values are presented as the median [IQR1–3]. ^*^Microhematuria was defined as urinary RBC ≥5/HPF. BMI, body mass index; RAAS, renin-angiotensin aldosterone system; SGLT2, sodium-glucose cotransporter 2; eGFR, estimated glomerular filtration rate; HDL, high density lipoprotein; LDL, low density lipoprotein.

**Supplementary Figure S1. Comparison of nephron level parameters across CKD stages in patients whose biopsy specimens contained ≥10 glomeruli and a cortical area of ≥4 mm^2^.**

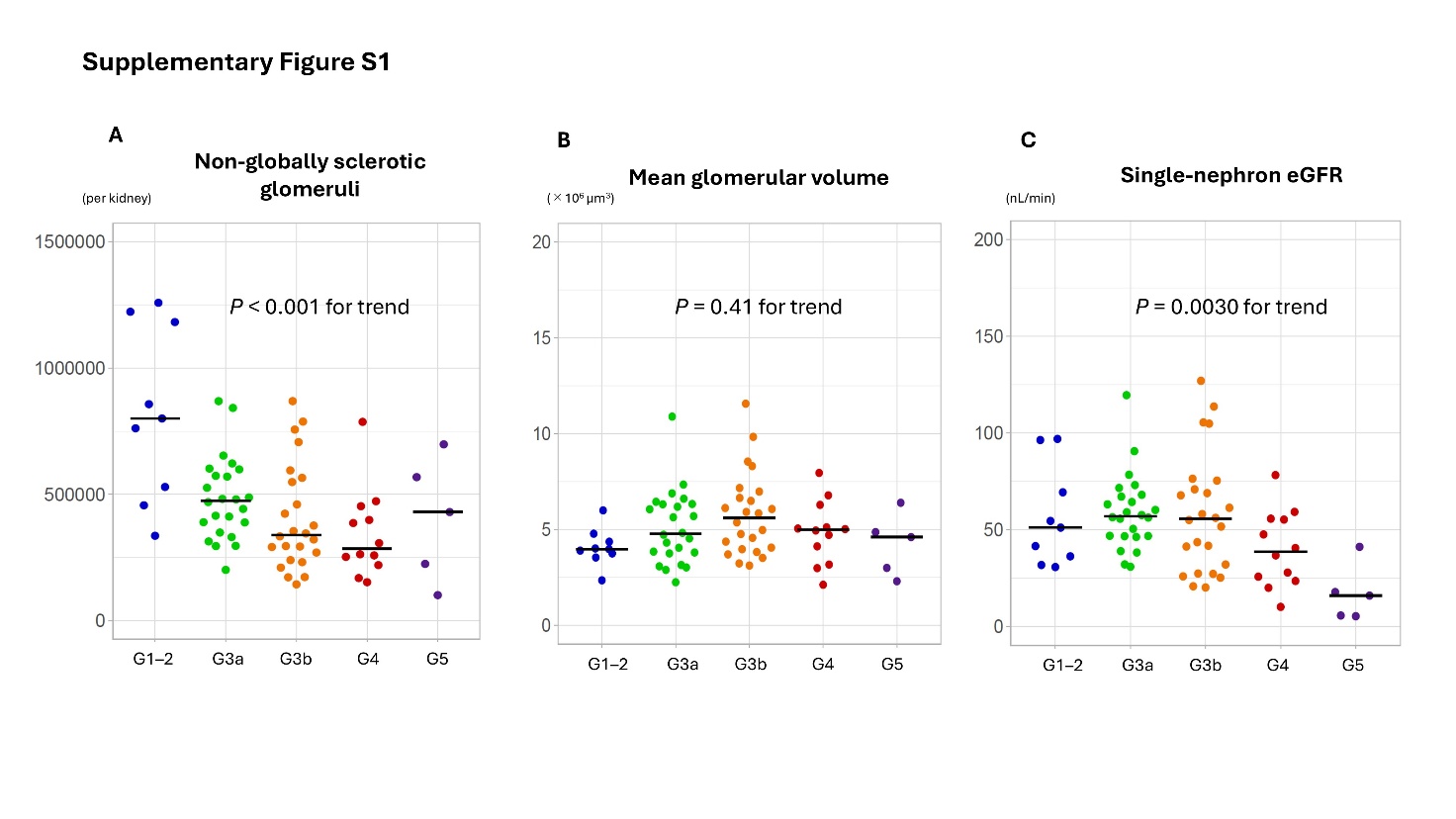

Nephron number per kidney (A), mean glomerular volume (B), and single-nephron eGFR (C) are shown at different CKD stages. CKD, chronic kidney disease; eGFR, estimated glomerular filtration rate.

**Supplementary Figure S2. Comparison of nephron-level parameters across CKD stages in patients treated with RAAS inhibitors.**

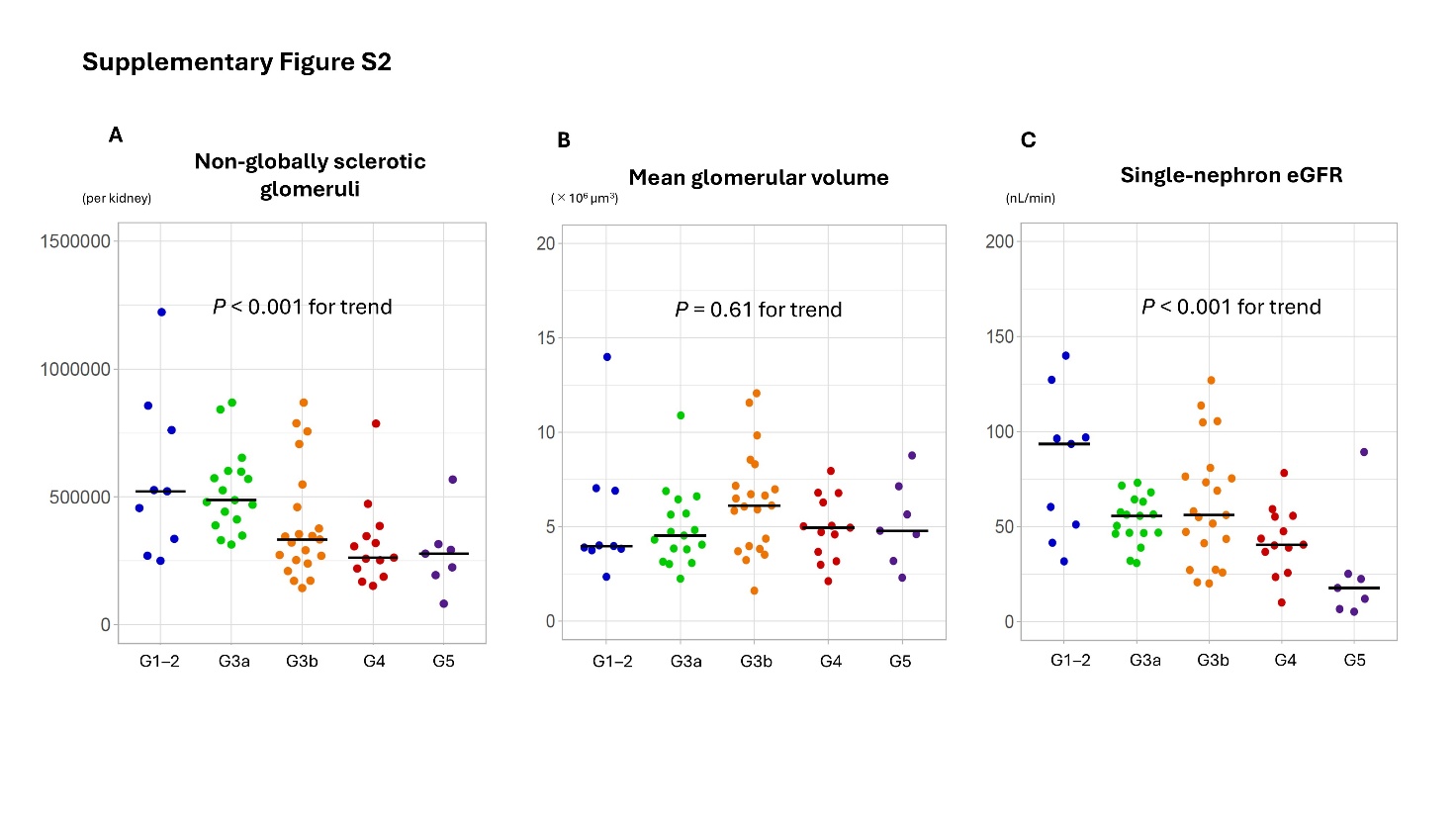

Nephron number per kidney (A), mean glomerular volume (B), and single-nephron eGFR (C) are shown at different CKD stages. CKD, chronic kidney disease; eGFR, estimated glomerular filtration rate.
